# Diagnostic Accuracy in Predicting the Clinical Diagnoses of Parkinson’s Disease, Parkinson’s Disease with Dementia and Dementia with Lewy Bodies Using Skin Biopsies Analyzed with Alpha-Synuclein Seed Amplification Assays

**DOI:** 10.1101/2025.05.21.25328106

**Authors:** Chelva Janarthanam, Christina D. Orrú, Anumantha G. Kanthasamy, Byron Caughey, Charles H. Adler, Holly A. Shill, David R. Shprecher, Andrew G. Hughson, Nan Zhang, Kewei Chen, Geidy E. Serrano, Thomas G. Beach

## Abstract

Skin biopsies of patients with parkinsonism, mild cognitive impairment or dementia might offer a simple and relatively non-invasive means to determine whether α-synuclein aggregates might be the underlying pathology. Accurate biomarkers are critically needed for Lewy body disease (LBD) clinical trials but currently there are none that have undergone full regulatory scrutiny.

We sought to simulate the rigor of a diagnostic study done with regulatory oversight in this study of the accuracy of skin biopsies processed by α-synuclein seeding amplification assays (SAA) in predicting the clinical diagnoses of Parkinson disease (PD), PD with dementia (PDD) and dementia with Lewy bodies (DLB). Our study design utilized parallel blinded performances of assays in two independent, experienced SAA assay laboratories. For control subjects without clinical LBD, we used a clinically heterogeneous set, simulating a cross-section of elderly, in comparison with previous studies that have mainly used cases and controls that have been prescreened to accentuate group differences.

Subjects clinically diagnosed with PD, PDD and DLB as well as those without a suspected LBD were recruited from three sites, including the Mayo Clinic Arizona, Barrow Neurological Institute and Banner Sun Health Research Institute (BSHRI), all located in metropolitan Phoenix, Arizona. The LBD group was designated as Group 1 while the non-LBD groups were divided between a Group 2 and a Group 3, based on the absence (Group 2) or presence (Group 3) of potential clinical risk factors for LBD, including mild cognitive impairment, dementia, REM sleep behavior disorder and hyposmia. Skin punch biopsies were collected from the posterior cervical area and three biopsies were analyzed by SAA between the two labs.

Sensitivity across assays ranged between 50.0% and 60.0% while specificity ranged between 69.6% and 100%. Comparisons of Group 1, with the highest clinical diagnostic confidence for the presence of an LBD, versus Group 2, with no clinical indicators of relevant abnormalities, produced the greatest specificities, between 77.3% and 100%, while sensitivities were less variable, ranging between 50% and 60% on group comparisons. Specificities were lower when Group 3 subjects, with potential LBD risk factors, were included in the calculations.

Pairwise agreement between biopsy assays ranged from excellent, with a kappa of 0.816, to moderate, between 0.47 and 0.67. Agreement may have been affected by differing protocols, as substrate and testing strategies were different between the two laboratories. Also, it might be possible that variability in α-synuclein seed concentrations within different skin biopsies might have contributed to differences in the outcome of the testing by the two labs. The greater agreement for Biopsy 3 was perhaps to be expected given that a single biopsy was analyzed by both labs and the homogenates were prepared by a single lab (Lab 2) and then shared between labs; homogenates for Biopsies 1 and 2 were separately prepared and separately assayed in Labs 1 and 2.

A separate analysis of subjects diagnosed with PDD or DLB indicated that skin SAA may have a greater sensitivity for clinically advanced LBD. Nine of these eleven subjects had at least one positive assay, for a sensitivity of 81.82%, substantially better than the 57.1% sensitivity for probable PD subjects without dementia. This suggests that skin SAA may have greater utility as a diagnostic and progression biomarker of Lewy body dementias, as compared with PD subjects without dementia.

## Introduction

Accurate diagnostic and progression biomarkers are essential for the optimization of clinical trials of disease-modifying therapeutics. As an example, the development of FDA-licensed PET amyloid methods that have been validated against autopsy histopathology have allowed a greatly increased ability to restrict the selection of Alzheimer’s disease (AD) clinical trial subjects to those who actually have significant concentrations of cortical amyloid plaques [1–5]. Currently, however, there are no biomarkers for Parkinson’s disease (PD) and other Lewy body disorders (LBDs) that have been adequately assessed against the autopsy gold standard for Lewy-type synucleinopathy (LTS) and none that have undergone full regulatory scrutiny. The US Food and Drug Administration (FDA) has historically allowed a less-stringent requirement for “laboratory-developed tests” (LDTs) as opposed to tests involving “devices” such as PET scans. To obtain Clinical Laboratory Improvement Amendments (CLIA) certification, for example, a new test must only show reproducibility; there is no requirement for accuracy in identification of the underlying pathogenic process or of a relationship with the clinical signs and symptoms. Recognizing this inadequacy, the FDA recently announced that it plans to bring LDTs under full scrutiny [6].

A major reason for the FDA’s concern [7] about LDTs has been their “high variability in performance…”. This has also been well documented for biopsy-based diagnostic tests for PD [8]. Colonic biopsies were the first that were widely pursued for PD diagnosis, quickly resulting in widely varying reports of diagnostic accuracy. Multicenter studies sponsored by the Michael J Fox Foundation eventually established that immunohistochemical (IHC) analysis of mucosal colonic biopsies were poorly sensitive and poorly specific for the clinical diagnosis of PD while full thickness evaluation of colon samples, only obtainable at autopsy, are highly specific for the defining brain autopsy neuropathology (9-10). This same problem now confuses the assessment of the utility of skin biopsies for PD, as the reported accuracy of these, whether analyzed using IHC or α-synuclein seeding amplification assays (SAA), has extensively varied between labs and methods, ranging from 24% to 98% [11–30].

A continuing and still largely unrecognized shortcoming of diagnostic test evaluations for PD and other synucleinopathies has been the persistent usage of an inadequate gold standard. The clinical diagnoses of Parkinson’s disease, dementia with Lewy bodies (DLB) and multiple system atrophy (MSA) are all inaccurate when compared to the autopsy diagnoses, especially for recognition of early disease stages, when the clinical diagnostic accuracy of early stage PD, for example, is documented as being only 71% or less [31–33]. For advanced DLB and MSA the respective figures are below even that [34–38]. The critical drawbacks of using “imperfect gold standards” have been repeatedly emphasized in the literature [39–44] and it is clear that testing the accuracy of a new diagnostic test against the clinical diagnosis can in fact lead to both underestimates and overestimates of its true (against autopsy) accuracy. Despite this problem, the clinical diagnoses of AD and PD are still routinely used to assess the accuracy of new biomarkers, without any admission of the uncertainty of this.

In the absence of strict regulatory oversight, we sought to partially simulate its rigor in conducting a test of the accuracy of skin biopsies processed by α-synuclein RT-QuIC, a form of SAA, in predicting the clinical diagnoses of PD, PDD and DLB. Key elements of our study design are the parallel performance of assays in two independent, experienced SAA assay laboratories, together with enforced blinding of the assay labs to subject diagnoses, as well as usage of cases and controls with comprehensive longitudinal clinical assessment by subspecialty neurologists. Additionally, we used a control subject set with heterogeneous clinical characteristics, to widely assess assay applicability across an elderly population, rather than using only narrowly-defined cases and controls that have been prescreened to accentuate the group differences.

Furthermore, although assessing accuracy against clinical diagnoses, we have applied our recently-developed method for expressing clinical diagnostic accuracy measures in terms of a range of theoretically possible values relative to autopsy-confirmed diagnoses [45].

## Methods

### Recruitment and Clinical Assessment

Subjects were recruited from three sites, including the Mayo Clinic Arizona, Barrow Neurological Institute and Banner Sun Health Research Institute (BSHRI), all located in metropolitan Phoenix, Arizona. Diagnostic criteria for PD were bradykinesia plus rest tremor and/or rigidity, with a positive response to dopaminergic therapy. For subjects enrolled in the Arizona Study of Arizona Study of Aging and Neurodegenerative Disorders (AZSAND) and Brain and Body Donation Program [46], additional definitions [31, 47] included: (1) probable PD (ProbPD): 2 of 3 cardinal signs (rest tremor, bradykinesia, cogwheel rigidity); no symptomatic cause; improvement when treated with dopaminergic medications and continued response if still being treated; or if lack of current response, then an explanation for why treatment was no longer working (such as inadequate dose because of side effects); (2) possible PD (PossPD): 2 of 3 cardinal signs, no symptomatic cause, symptoms or signs present for ≤5 years, dopaminergic treatment had not been tried, or an adequate trial had not clearly occurred (i.e., too low a dose, side effects that limited the therapeutic dose); (3) suspect PD (SuspPD): only 1 sign of PD, either rest tremor of the jaw or limbs or bradykinesia with no symptomatic cause; (4) parkinsonism not otherwise specified (ParkNOS): parkinsonism without response to an adequate dose of dopaminergic medication or disease duration of >5 years; parkinsonism that had not been treated or been given an adequate trial of dopaminergic treatment; or parkinsonism that seemed to have another etiology including an unclear neurodegenerative condition, dementia with parkinsonian features, or secondary parkinsonism.

For DLB, published diagnostic criteria from the Dementia with Lewy Bodies Consortium were used, incorporating minimal combinations of core clinical features including visual hallucinations, parkinsonism, cognitive or attention fluctuations and clinical features consistent with REM sleep behavior disorder [48].

Clinical assessment included, at minimum, the Clinical Dementia Rating (CDR) Scale [49,50], Mini Mental State Examination (MMSE), Montreal Cognitive Assessment (MoCA) [51–53], Unified Parkinson’s Disease Rating Scale (UPDRS)[54] and Hoehn and Yahr Scale [55]. For most subjects (95/105; 90.5%), the full neuropsychological and movement disorders battery that is part of the AZSAND [46] was used to establish the relevant clinical history, presence or absence of dementia and the degree of cognitive impairment. Mild cognitive impairment and dementia were minimally defined as having global CDR scores of 0.5 and 1 or greater, respectively, while for AZSAND subjects this was decided by clinical consensus conferences reviewing the full test batteries by participating neuropsychologists and cognitive/behavioral neurologists.

Aside from clinical diagnosis, other inclusion criteria were: 1. Males or females ≥ 40 years of age. 2. Willingness to undergo the required clinical assessments and biopsies. Exclusion criteria were: 1. Are being treated with chemotherapy for cancer. 2. Are known to have a structural brain lesion or other CNS disease process or symptoms. 3. Have a clinically significant infectious disease, such as HIV, hepatitis or prion disease. 4. Have suspected encephalopathy due to alcoholism or end-stage liver disease. 5. Have a bleeding diathesis, are on anticoagulant therapy, or have severe medical illness. 6. Have skin disease in the posterior cervical area.

For statistical analyses, subjects who were clinically diagnosed with PD, PDD or DLB were designated as Group 1, while Group 2 subjects were defined as those who were cognitively unimpaired without any signs of parkinsonism, and Group 3 subjects were those with potential risk factors for underlying Lewy body disease (LBD), including mild cognitive impairment (MCI), dementia, AZSAND clinical diagnoses of “suspect PD”, “possible PD”, “parkinsonism NOS”, a clinical history consistent with REM sleep behavior disorder (RBD), or hyposmia, defined as a University of Pennsylvania Smell Identification Test (UPSIT) score of 20 or less.

Clinical data used for subject classification was obtained from two timepoints, including both the assessment done closest to biopsy date, within one year before or after biopsy, and the final assessment, done two or more years after biopsy. Diagnostic accuracy figures calculated included sensitivity, specificity, positive and negative predictive values, and positive and negative likelihood ratios. Agreement between assays was assessed with Cohen’s Pairwise Kappa.

### Skin Biopsies

Six posterior cervical skin punch biopsies were performed on all subjects with few exceptions. Skin punch biopsies were obtained by neurologists, research nurses, physicians’ assistants or nurse practitioners using methods as previously described for the Michael J Fox Foundation S4 study [56], with the disposable Miltex Biopsy Punch, 3 mm internal diameter (obtained from Electron Microscopy Science, catalogue number 69038-03), after wiping the skin with an alcohol swab and local anesthetic injection, at approximately the C7-C8 level. Bilateral biopsies were taken, each set of 3 located approximately 3 cm from the midline (Fig. 1, Supplementary. Methods). The posterior cervical location was chosen as it has been found to have higher densities of Lewy-type synucleinopathy (LTS) than more distal locations including the extremities, by AZSAND autopsy data, data from the S4 Study [11] and in a study by Donadio et al of the relative LTS positivity in different sites [57].

Biopsied tissue was immediately placed between two sponges in standard plastic paraffin-embedding cassettes, immersed in 50 ml of cold sterile normal saline and transported on ice the same day to the Civin Laboratory for Neuropathology at Banner Sun Health Research Institute, which served as the pathology core for this project. The biopsies were there removed from the cassettes, placed into microcentrifuge tubes, rapidly frozen on dry ice and stored in a −80°C ultralow temperature freezer. A single biopsy from each subject was subsequently fixed in 10% formalin, paraffin embedded, sectioned and stained with H & E, and examined for adequacy by a pathologist (TGB). Other biopsies were later shipped on dry ice by air courier service to the three RT-QuIC assay laboratories. Coding of biopsies was done to keep the assay labs blinded to clinical diagnosis, with the codes kept at the project’s statistical core at the Arizona Mayo Clinic.

### RT-QuIC Analysis of Skin Biopsies

Two independent laboratories performed RT-QuIC assays on the frozen biopsies. These are designated here as: Lab 1, headed by Byron Caughey, PhD and located in Hamilton, Montana at the Laboratory of Neurological Infections and Immunity of the National Institute for Allergies and Infectious Diseases; Lab 2, headed by Anumantha Kanthasamy, PhD, located in Athens, Georgia at the University of Georgia.

Labs 1 and 2 were initially given one biopsy each, designated here as Biopsy 1 and Biopsy 2, that they then processed according to their own protocols. Following this, Biopsy 3 was given to Lab 2, where it was homogenized as for their protocol for Biopsy 2 and aliquots sent to Lab 1; these homogenates were then processed by each lab for RT-QuIC in the same way as done for Biopsies 1 and 2.

Some initial experiments were done by Lab 2 to tailor their homogenization and assay conditions to the biopsies received,and assess for possible interference with skin SAA by keratin (see Supplementary Methods).

### Homogenization protocols

At Lab 1, biopsies were thawed and washed three times in TBS. The tissues were minced in 10% (w/v) TBS containing 2 mM CaCl2 and 0.25% (w/v) collagenase A for one-minute maximum speed using the Mini-Beadbeater. Next, samples were incubated at 37°C and 400 rpm shaking for 4 hours, then homogenized a second time with the Mini-Beadbeater, again for one minute at maximum speed. The final homogenate was then centrifuged at 2000 g for 2 minutes and the supernatant distributed in single use aliquots for storage at - 80°C.

At Lab 2, biopsies were weighed, sliced into smaller pieces and washed with 1X TBS until no visible blood contamination remained. Tissue lysis buffer (2 mM CaCl₂, 0.25% [w/v] collagenase A; Millipore Sigma, Darmstadt, Germany; Cat# 10103578001 in 1× TBS) was added to achieve a final concentration of 10% (w/v) in a 1.5-mL Eppendorf tube. Samples were incubated at 37°C in a thermomixer shaking at 350 rpm for 4 h.

Following incubation, the samples were transferred to 1.5-mL RINO tubes containing 0.5-mm zirconium beads and homogenized using a bullet blender for five 1-min cycles, with a 1-min rest on ice between each cycle. The homogenates were then centrifuged at 500 × g, and the supernatants were collected and aliquoted into 10-μL volumes. The 10% homogenates were stored at −80°C until further use.

### Assay protocols

At Lab 1, clear-bottom 96-well plates (Thermo Fisher Scientific, Waltham, MA, USA; Cat# 1256672) were pre-loaded with six 800-µm silica beads (OPS Diagnostics, Lebanon, NJ, USA; Cat# 80020002) per well. The reaction mixture contained 0.1 mg/mL (measured via NanoDrop) of 100 kDa (Pall) filtered in-house purified mutant αSyn K23Q-αSyn monomer [58] (Accession No. NM_000345.3), 10 µM ThT, 40 mM phosphate buffer (pH 8.0), and 170 mM NaCl (see also Supplementary Methods). Each well received 98 µL of the reaction mixture, followed by 2 µL of 1:100 dilution from 10% skin homogenates: final skin tissue dilution per quadruplicate reactions was 1:1000. The plates were sealed with a plate sealer (Thermo Fisher Scientific, Waltham, MA, USA; Cat# 235307) and incubated in a BMG FLUOstar Omega (BMG Labtech, Ortenberg, Germany) at 42°C with 400 rpm double orbital shaking for 1 min followed by 1-min rest with ThT fluorescence readings (445-nm excitation and 480-nm emission wavelengths), every 45 min.

At Lab 2, 96-well clear-bottom plates (Thermo Fisher Scientific, Waltham, MA, USA; Cat# 1256672) were loaded with six 800-µm silica beads (OPS Diagnostics, Lebanon, NJ, USA; Cat# 80020002) in each well. In-house purified human WT-αSyn monomer [59] was thawed from −80°C and centrifuge-filtered (100 kDa) before use (see also Supplementary Methods). The reaction mixture contained 0.1 mg/mL filtered αSyn monomer (as measured by the NanoDrop), 10 µM ThT, 40 mM phosphate buffer (pH 8.0), 0.00125% SDS, and 40 mM NaCl. Next, 98 µL of the reaction mixture was loaded into each well of the plate, followed by 2 µL of 1:10 dilution from 10% skin homogenates; final dilution of skin samples was 1:100. The plates were sealed with a plate sealer (Thermo Fisher Scientific, Waltham, MA, USA; Cat# 235307) and incubated in the BMG FLUOstar Omega (BMG Labtech, Ortenberg, Germany) at 42°C. The reader settings were as follows: double orbital shaking at 400 rpm for 1 min followed by 1-min resting incubation between shaking cycles; gain was set to 1300; ThT fluorescence was measured every 30 min for a minimum of 42 h using 445-nm excitation and 480-nm emission wavelengths. The samples were run as quadruplicates. The kinetic parameters evaluated were protein aggregation rate (PAR), fluorescence maximum (Fmax) and hit rate (number of replicates crossing threshold). The threshold line used to calculate PAR and hit rate was defined as the mean plus 10 standard deviations (µ+10σ), calculated from the first ten readings of all samples.

### Criteria for positive vs negative assay results

For both labs, the primary kinetic criterion used to label a sample positive was the number of replicate reactions crossing the designated fluorescence threshold. For Lab 1, the thioflavin T (ThT) fluorescence threshold was defined as 10% of the maximum value on each plate within an assay incubation time of 40hs. For Lab 2, the threshold of 5,000 RFU was derived as µ+10σ, calculated from the first 10 readings of all samples (with 4 unblinded controls).

For both Labs, samples were tested in quadruplicate and classified as 4/4, 3/4, 2/4, 1/4, or 0/4 well hits; when an SAA-tested sample was positive in 4/4 or 3/4, it was defined as overall positive without retesting. For Lab 1, samples scoring 2/4 were retested, whereby retested samples scoring ≥ 2/4 were defined as positive; otherwise, samples scoring 1/4 or 0/4 were considered negative. At Lab 2 this same procedure was followed but retesting was done with additional dilutions, to reduce chances of negative results occurring due to samples having high concentrations of seeds, or high protein or lipid levels, as both situations could inhibit seeding signals.

## Results

The completeness of biopsies was assessed on H & E slides for one biopsy from each subject; all biopsies were judged adequate in that they included at least the dermis layer, where innervation is concentrated, while the majority included all three layers, i.e. epidermis, dermis and hypodermis. Summary data for study subjects are given in Tables 1 and 2. All subjects were non-Hispanic and white/Caucasian except for one Asian subject. At the time of biopsy, Group 1 subjects included 26 with PD and 6 with DLB; 8 of the PD subjects had MCI and 3 had dementia (PDD); their mean symptom duration was 8.1 years (SD 4.7). Group 3 subjects included 15 with parkinsonism not meeting criteria for probable PD (possible PD, suspect PD or PD-NOS, see Methods), 18 with MCI and 7 with dementia. Group 1 subjects significantly differed from Groups 2 and/or 3 in age (group 1 was youngest), sex distribution (more males in Group 1), proportion with RBD (more RBD in Group 1) and UPDRS motor score (higher scores in Group 1), but were not significantly different in their MoCA scores or years of education.

**Table 1.** Clinical diagnoses by group at initial diagnosis. Group diagnoses are non-exclusive. Poss = Possible PD, not meeting full PD criteria; Susp = Suspect PD, not meeting full PD criteria; Park NOS = non-specific parkinsonism, Not Otherwise Specified. The dementia category includes probable Alzheimer’s disease, vascular dementia and dementia NOS. N/A = Not available or not present.

| Group | Group 1 (n = 32) | Group 2 (n = 22) | Group 3 (n = 51) |
| --- | --- | --- | --- |
| PD Diagnosis (all) | 26 (81.2%) | N/A | N/A |
| PD MCI Diagnosis (subset) | 8 (25.0%) | N/A | N/A |
| PDD Diagnosis (subset) | 3 (9.37%) | N/A | N/A |
| DLB Diagnosis | 6 (18.7%) | N/A | N/A |
| Poss/Susp/ParkNOS | N/A | N/A | 15 (29.4%) |
| MCI | N/A | N/A | 18 (35.3%) |
| Dementia | N/A | N/A | 7 (13.7%) |

**Table 2.** Demographics and quantitative clinical measures by initial group diagnoses. Hyposmia is defined as having a UPDRS score of 20 or less.

| Group | Group 1 (n = 32) | Group 2 (n = 22) | Group 3 (n = 51) |
| --- | --- | --- | --- |
| Age at Biopsy<sup>1</sup> | 72.7 (SD 6.2) | 77.7 (SD 7.8) | 79.7 (SD 8.5) |
| Sex<sup>2</sup> | 25 males (78.1%) | 5 males (22.7%) | 30 males (58.8%) |
| Yrs Education | 16.0 (SD 2.6) | 15.6 (SD 2.6) | 18.4 (SD 13.7) |
| Signs/Symptoms | 8.1 (SD 4.7) | N/A | N/A |
| RBD Clinical Dx<sup>3</sup> | 17 (53.1%) | N/A | 9 (17.6%) |
| Hyposmia | 19 (59.4%) | N/A | 28 (54.9%) |
| UPDRS III Score<sup>4</sup> | 29.10 (SD 19.4) | 2.6 (SD 3.3) | 7.1 (SD 8.1) |
| MoCA Score | 20.6 (SD 6.3) | 22.7 (SD 3.9) | 19.7 (SD 6.1) |

1. Group 1 significantly younger than Group 2 (p = 0.01) and Group 3 (p < 0.001). 2. Group 1 has significantly more males than Group 2 (p = 0.0001). 3. Group 1 has significantly more RBD than Group 2 or 3 (p < 0.01). 4. Group 1 has significantly greater UPDRS motor scores than Groups 2 or 3 (p < 0.0001).

Some subjects had changed diagnoses (data available on request) between the time of their biopsy (2021 or 2022) and the study end (May 1, 2025 or time of dropout or death). One Group 1 subject with an initial diagnosis of probable PD was later classified as Group 3 after being clinically diagnosed as progressive supranuclear palsy. Five Group 2 subjects without any significant diagnoses were later classified as Group 3 due to subsequent diagnoses of one or more of MCI, vascular dementia, suspected PD or hyposmia. Two Group 3 subjects were later classified as Group 2 after reverting to having no significant diagnoses. Two Group 3 subjects were later classified as Group 1 after being diagnosed with probable PD.

Tables 3, 4 and 5 below give sensitivity and specificity for all biopsies and assays. Positive and negative results for all assays are available on request. The Supplementary Data File gives a more detailed summary of statistical characterizations, including sensitivity, specificity, positive and negative predictive value (PPV, NPV), overall accuracy and positive and negative likelihood ratios (LR). Diagnostic accuracy was calculated for each biopsy and lab, using both diagnosis at time of biopsy and at study end, and under different conditions, including comparisons of Group 1 versus Groups 2 and 3 combined, Group 1 versus Group 2 alone, and Group 1 versus Group 3 alone. For all comparisons, a biopsy was considered positive if one or more assays by either lab was rated as positive and considered negative if all assays had a negative result. Under this rule, sensitivity across assays ranged between 50.0% and 60.0% while specificity ranged between 69.6% and 100%. The greatest specificities were achieved in the comparisons of Group 1 versus Group 2, between 77.3% and 100%, while sensitivities uniformly ranged between 50% and 60%. Specificities were somewhat lower when Group 3 subjects were included in the calculations, as expected given their possession of potential risk factors for harboring clinically undeclared LBD. Generally, neither sensitivity nor specificity changed substantially between initial and followup diagnoses so Tables 3-5 below show sensitivities and specificities only for the initial diagnoses; see Supplementary Data File for full statistical summaries for subjects clinically classified at both initial and followup clinical assessments.

**Table 3.**
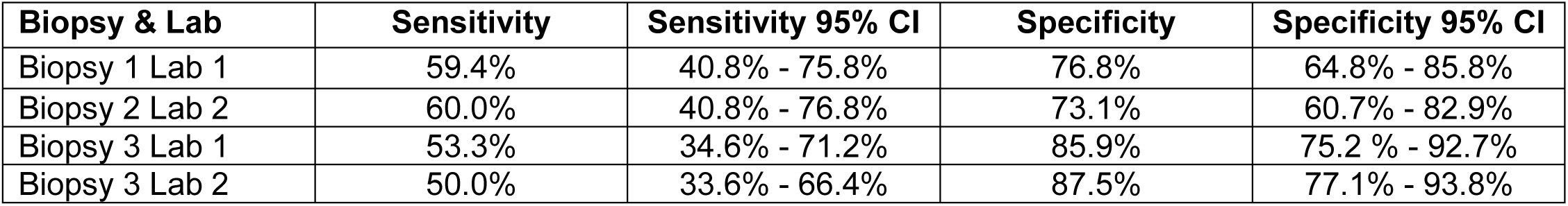
Sensitivity and specificity for all assay results, using initial diagnosis and including all three diagnostic groups. Group 1 is defined as the disease state while Groups 2 and 3 are defined as non-disease.

**Table 4.** Sensitivity and specificity for all assay results, using initial diagnosis. Group 1 is defined as the disease state while Group 2 is defined as non-disease.

| Biopsy & Lab | Sensitivity | Sensitivity 95% CI | Specificity | Specificity 95% CI |
| --- | --- | --- | --- | --- |
| Biopsy 1 Lab 1 | 59.4% | 40.8% - 75.8% | 77.3% | 54.2% - 91.3% |
| Biopsy 2 Lab 2 | 60.0% | 40.8% - 76.8% | 81.0% | 57.4% - 93.7% |
| Biopsy 3 Lab 1 | 53.3% | 34.6% - 71.2% | 90.9% | 69.4% - 98.4% |
| Biopsy 3 Lab 2 | 50.0% | 33.6% - 66.4% | 100% | 80.8% - 100.0% |

**Table 5.** Sensitivity and specificity for all assay results, using initial diagnosis. Group 1 is defined as the disease state while Group 3 is defined as non-disease.

| Biopsy & Lab | Sensitivity | Sensitivity 95% CI | Specificity | Specificity 95% CI |
| --- | --- | --- | --- | --- |
| Biopsy 1 Lab 1 | 59.4% | 40.8% - 75.8% | 76.6% | 61.6% - 87.2% |
| Biopsy 2 Lab 2 | 60.0% | 40.8% - 76.8% | 69.6% | 54.1% - 81.8% |
| Biopsy 3 Lab 1 | 53.3% | 34.6% - 71.2% | 83.7% | 69.8% - 92.2% |
| Biopsy 3 Lab 2 | 50.0% | 33.6% - 66.4% | 82.4% | 68.6% - 91.1% |

We separately analyzed the sensitivity of SAA for subjects diagnosed with PDD or DLB. As there were only 5 cases of PDD and 6 with DLB, we combined them for this calculation. Nine of these eleven subjects had at least one positive assay, for a sensitivity of 81.82%, substantially better than the 57.1% sensitivity for probable PD subjects without dementia, of whom only 12/21 were SAA positive.

Results agreement between biopsies was first assessed by calculating the intraclass correlation coefficient (ICC). Using a one-way model (assuming the subjects were randomly selected) and absolute agreement, the 95% confidence intervals were calculated across the four biopsies. The overall ICC is 0.605 (95% CI: 0.511 – 0.694), indicating a moderate reliability across assay results. Using only single initial diagnostic groups, the ICC in Group 1 is the highest at 0.73 (95% CI: 0.586 – 0.847), while it was lowest for Group 2 at 0.255 (95%

CI: 0.049 – 0.522) and between the two for Group 3, at 0.454 (95% CI: 0.305 – 0.608). Further, Cohen’s kappa was calculated for the pairwise agreement between any two biopsies and the results are shown in Table 6 below. There was excellent agreement between the two labs for Biopsy 3 (kappa = 0.816) while all the other paired results showed moderate to substantial agreement (0.47-0.67).

**Table 6.** Pairwise Cohen’s kappa between all pairs of assays, at initial diagnosis.

| Biopsy & Lab | Biopsy 1 Lab 1 | Biopsy 2 Lab 2 | Biopsy 3 Lab 1 |
| --- | --- | --- | --- |
| Biopsy 2 Lab 2 | 0.548 |  |  |
| Biopsy 3 Lab 1 | 0.669 | 0.516 |  |
| Biopsy 3 Lab 2 | 0.645 | 0.470 | 0.816 |

As the diagnostic accuracy for the clinical diagnosis of probable PD at AZSAND, validated against autopsy neuropathology, is 70.6% in subjects with a disease duration less than 5 years and 89.1% with a disease duration equal to or greater than 5 years disease duration [31] and 7 of the 26 Probable PD cases (26.9%) in this study had a disease duration less than 5 years, it might be roughly estimated that the accuracy (both sensitivity and specificity) of the clinical diagnosis of the Probable PD group in this study might have been about 80%. Using the method we have developed for interpreting biomarker test results in the absence of the autopsy gold standard [45], and assuming that AZSAND diagnostic accuracy across our entire Group 1 cases might be similar, despite including 6 DLB cases, we estimate that our skin RT-QuIC results obtained here would have a likely true sensitivity for identifying Group 1 cases as ranging between approximately 40% and 54% and the likely true specificity of the biopsy assays for identifying non-Group 1 cases would range between 64% and 90% (see Figure 1 of Supplementary Data File), although we cannot rule out the possibility that the true specificity of skin SAA, against autopsy, may exceed that of the clinical diagnosis of PD/LBD, as there have been no published definitive, autopsy-validated studies of this. If so, then our calculated ranges may underestimate the true ranges.

## Discussion

Skin biopsies of patients with parkinsonism, mild cognitive impairment or dementia might offer a simple and relatively non-invasive means to determine whether α-synuclein pathology might be the underlying pathology. However, whether analyzed using either IHC or α-synuclein seeding amplification assays (SAA), the reported accuracy for diagnosing PD has extensively varied between labs and methods, ranging from 24% to 98% [11–30]. Accurate diagnostic and progression biomarkers are critically needed for LBD clinical trials but currently there are no biomarkers for LBDs that have been adequately validated against the autopsy gold standard and none that have undergone full regulatory scrutiny.

In the absence of sufficiently strict regulatory oversight, we sought to partially simulate its rigor in this study of the accuracy of skin biopsies processed by SAA in predicting the clinical diagnoses of PD, PDD and DLB. Our study design utilized the parallel performance of assays in two independent, experienced SAA assay laboratories, together with enforced blinding of the assay labs to subject diagnoses. Additionally, we used mostly cases and controls with comprehensive longitudinal clinical assessment by subspecialty neurologists and neuropsychologists. Furthermore, our usage of a control subject set with heterogeneous clinical characteristics is more widely applicable across an elderly population, in comparison with previous studies that have mainly used cases and controls that have been prescreened to accentuate group differences.

Sensitivity across assays ranged between 50.0% and 60.0% while specificity ranged between 61.5% and 100%. Comparisons of Group 1, with the highest clinical diagnostic confidence for the presence of an LBD, versus Group 2, with no clinical indicators of relevant abnormalities, produced the greatest specificities, between 77.3% and 100%, while sensitivities were less variable, ranging between 50% and 60%. Specificities were lower when Group 3 subjects, with potential LBD risk factors including MCI, dementia, RBD and hyposmia, were included in the calculations.

Our separate analysis of subjects diagnosed with PDD or DLB, although a small sample of only 11 subjects, indicated that skin SAA may have a greater sensitivity for clinically advanced LBD. Nine of these eleven subjects had at least one positive assay, for a sensitivity of 81.82%, substantially better than the 57.1% sensitivity for probable PD subjects without dementia, where 12/21 were SAA positive. This suggests that skin SAA may have greater utility as a diagnostic and progression biomarker of Lewy body dementias, as compared with PD subjects without dementia. Larger numbers of subjects are needed to confirm or refute this.

The pairwise agreement between biopsy assays was excellent, with a kappa of 0.816, for Biopsy 3, while all other paired results had moderate to substantial agreement, ranging between 0.47 and 0.67. Agreement may have been affected by differing protocols, substrate and testing strategies that were different between the two laboratories. Also, it might be possible that variability in α-synuclein seed concentrations within different skin biopsies might be contributing to differences in the outcome of the testing by the two labs. The greater agreement for Biopsy 3 was perhaps to be expected given that a single biopsy was analyzed by both labs and the homogenates were prepared by a single lab (Lab 2) and then shared between labs; homogenates for Biopsies 1 and 2 were separately prepared and separately assayed in Labs 1 and 2.

Importantly, for the first time in any biomarker report that we are aware of, and using a method that we have recently published [45], we have calculated estimates of the true accuracy of our assay results, had they been done on autopsy-confirmed cases.

## Data Availability

All data produced in the present work are available on reasonable request to the authors.

