## Supplementary Data File 05-21-25 for "Diagnostic Accuracy in Predicting the Clinical Diagnoses of Parkinson’s Disease, Parkinson’s Disease with Dementia and Dementia with Lewy Bodies Using Skin Biopsies Analyzed with Alpha-Synuclein Seed Amplification Assays"

**Table 1.** Detailed results analysis for Labs and Biopsies at initial diagnosis, with Group 1 defined as Disease and Groups 2 and 3 defined as Non-Disease (n = 105).

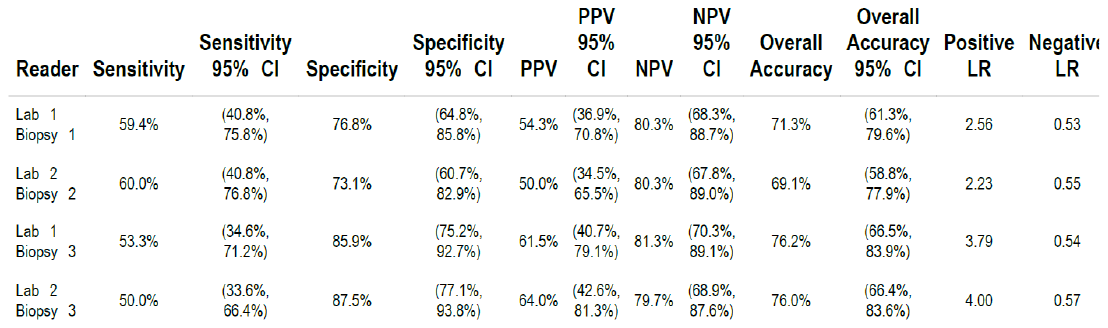

**Table 2.** Detailed results analysis for Labs and Biopsies at followup diagnosis, with Group 1 defined as Disease and Groups 2 and 3 defined as Non-Disease (n = 105).

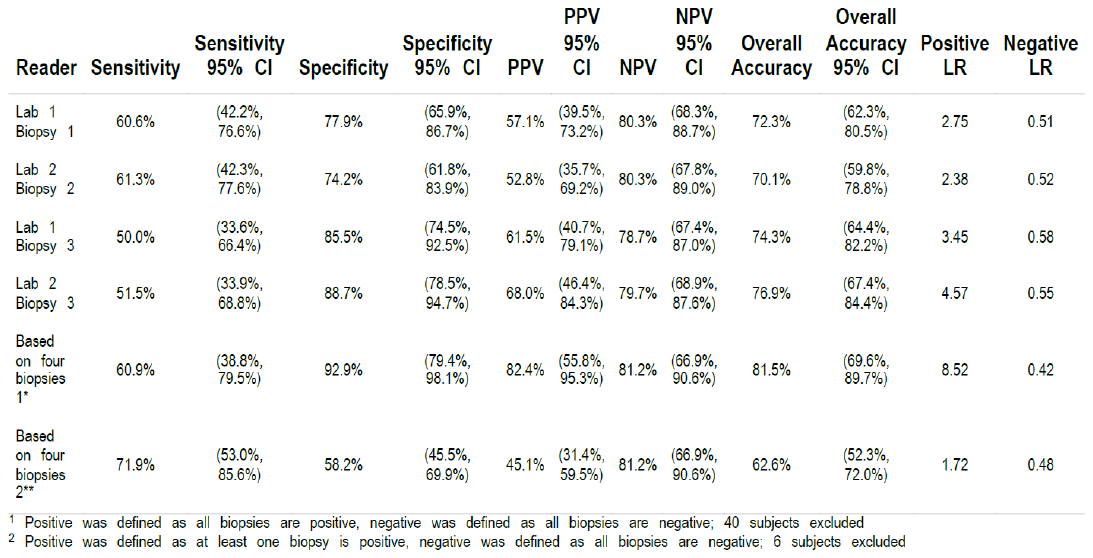

**Table 3.** Detailed results analysis for Labs and Biopsies at initial diagnosis, with Group 1 defined as Disease and Group 2 defined as Non-Disease (n = 54).

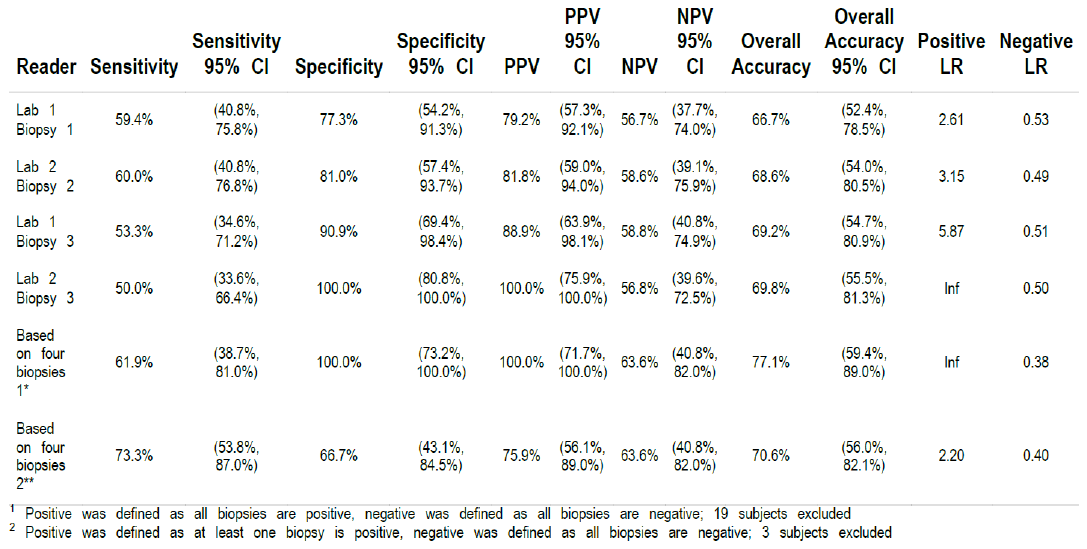

**Table 4.** Detailed results analysis for Labs and Biopsies at followup diagnosis, with Group 1 defined as Disease and Group 2 defined as Non-Disease (n = 53).

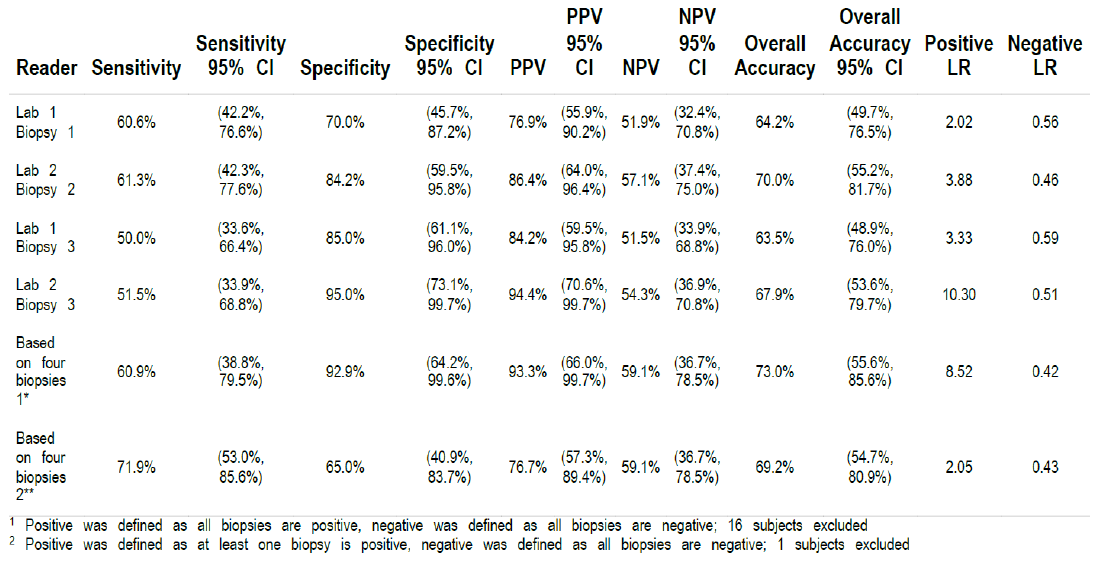

**Table 5.** Detailed results analysis for Labs and Biopsies at initial diagnosis, with Group 1 defined as Disease and Group 3 defined as Non-Disease (n = 83).

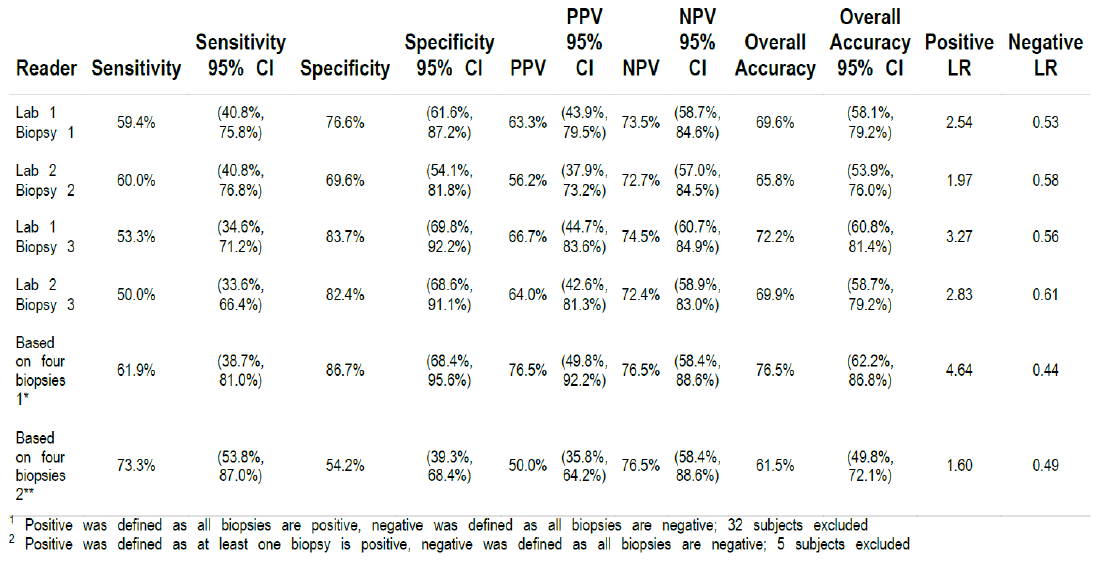

**Table 6.** Detailed results analysis for Labs and Biopsies at followup diagnosis, with Group 1 defined as Disease and Group 3 defined as Non-Disease (n = 85).

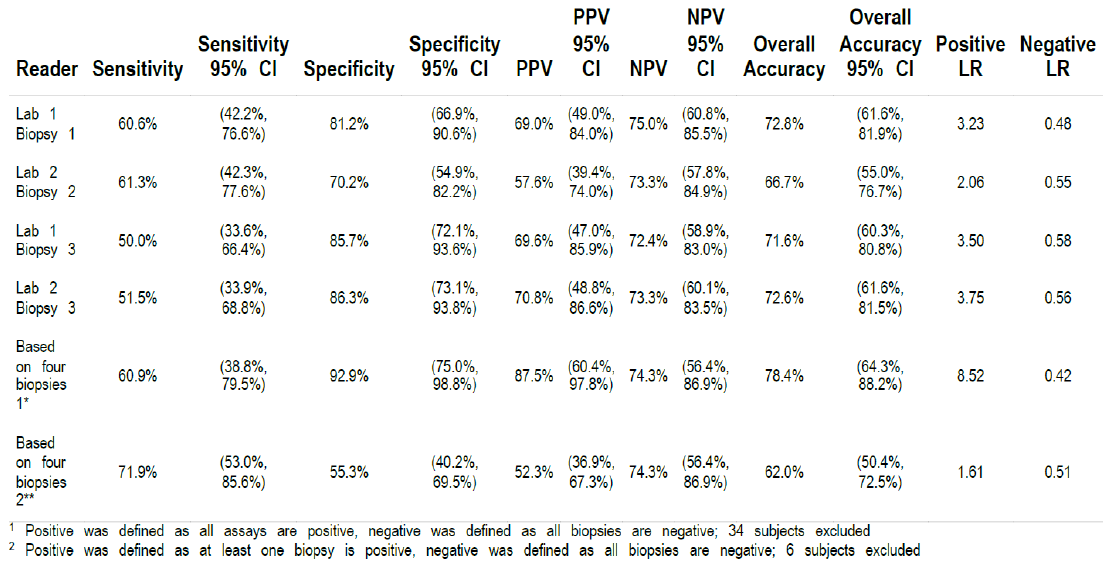

**Figure 1.** Probabilistic range of true sensitivity and specificity of observed biomarker values, assuming true diagnostic sensitivity and specificity of the clinical diagnosis at AZSAND is 80% for each. The ranges are depicted as arising from the 95% confidence intervals for the observed sensitivity and specificity in Tables 3, 4 and 5 from the main manuscript. Here the likely true sensitivity of the biopsy assays for identifying Group 1 cases are shown as ranging between approximately 40% to 54% and the likely true specificity of the biopsy assays for identifying non-Group 1 cases as ranging between 64% and 90%. The red dashed line represents a perfect biomarker, i.e. a biomarker that equals the accuracy of the autopsy diagnosis From Zhang N et al. Interpreting Biomarker Test Results for Alzheimer’s Disease, Parkinson’s Disease and Other Neurodegenerative Diseases Without the Autopsy Gold Standard. medrxiv doi: <https://doi.org/10.1101/2025.04.23.25326286>.

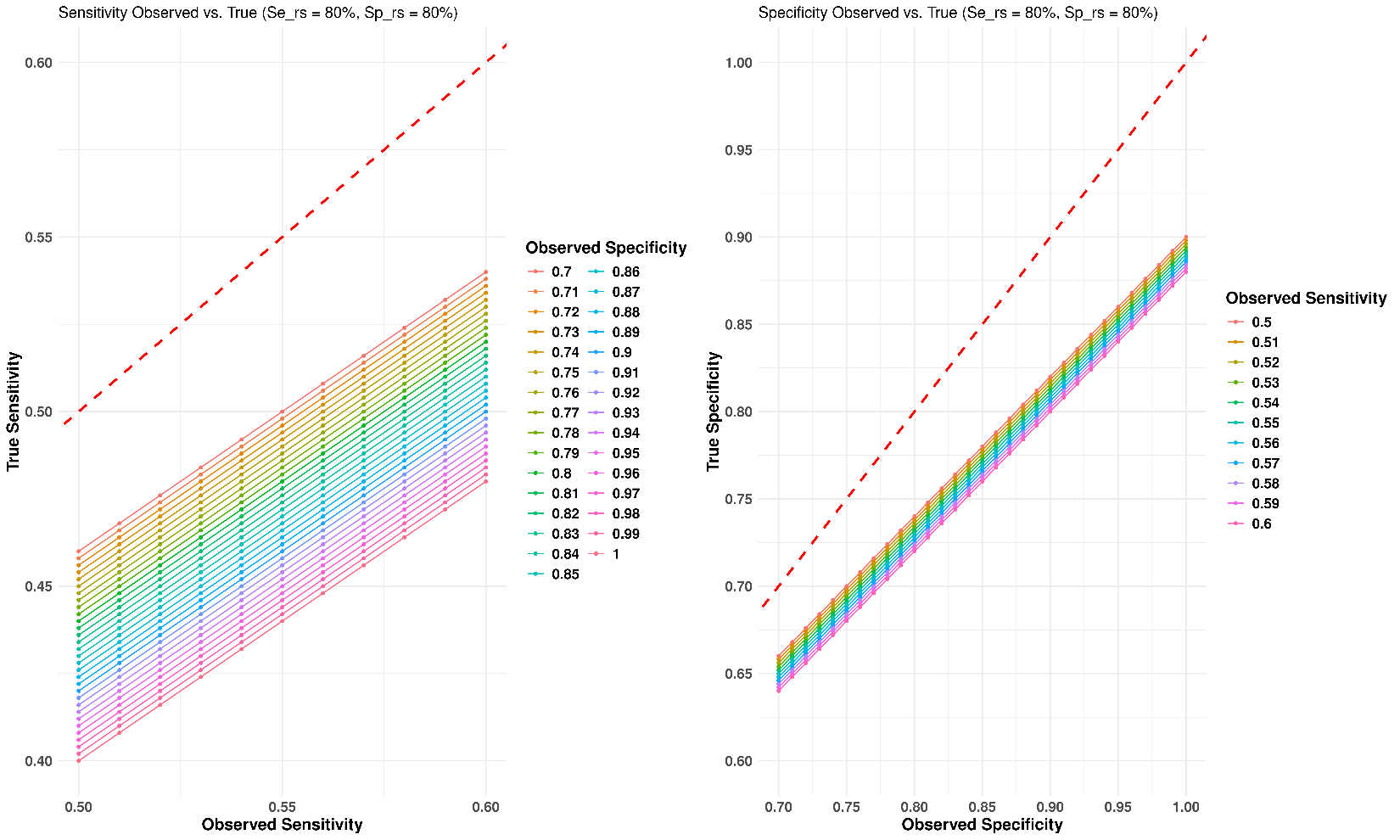
