## Supplementary Methods 05-20-25 for "Diagnostic Accuracy in Predicting the Clinical Diagnoses of Parkinson’s Disease, Parkinson’s Disease with Dementia and Dementia with Lewy Bodies Using Skin Biopsies Analyzed with Alpha-Synuclein Seed Amplification Assays"

**Figure 1.** Topographic location of skin biopsies, at the base of the back of the neck, on either side of the spinal levels C7 and C8. Three skin punches were taken from each subject, on either side of the midline, for a total of 6 biopsies.

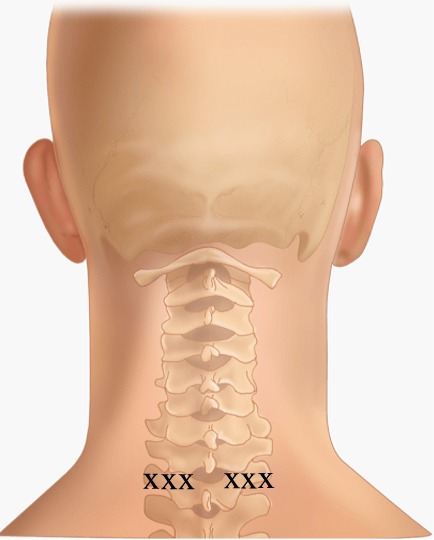

**Lab 1 K23Q recombinant α-Syn purification**

Using Q5 Site-Directed Mutagenesis (NEB) and the primers CACACCCTGTTGGGTTTTCTCAG and CAGAAGCAGCAGGAAAGAC, we engineered the K23Q mutation into the wild type αSyn sequence (Accession No. NM_000345.3) in a pET28 vector with an N-terminal His-tag (EMD Biosciences), as previously described [ref 58 in main manuscript]. We transformed the plasmid into BL21(DE3) Escherichia coli (EMD Biosciences) for protein expression. Notably, protein was purified either from the periplasmic space or from whole cell lysates interchangeably. Sensitivity and specificity of proteins isolated using both techniques were compared and no significant difference was observed in their RT-QuIC performance Recombinant αSyn was purified from the periplasmic space as previously described. . Whole cell lysate isolates were obtained following an overnight auto-induction expression. The 1L Luria broth BL21(DE3) cell suspension was split into 4x250mL conical vessels, spun at 3273xg for 12min, and cell pellets were frozen at -80 °C for 20min. They were then resuspended in 30ml of cold PBS per vessel. Next, each suspension was transferred into a 50ml falcon tube,probe sonicated in an ice bath (4 x 45sec with 15sec rests) at a power setting of 45% (Ultrasonic) and each tube was boiled for 20min and centrifuged at 9000xg for 60min. Supernatants were combined, 50ml of Buffer A (20mM Tris pH7.4) were added and the supernatant was filtered (0.22 μm vacuum filter) and loaded onto a 5mL Ni-NTA column (Cytiva 17525501). We collected the middle section of the elution peak and added 20mL of Buffer A before moving onto a 5ml Q-HP (Cytiva 17115401) column using pre-chilled Buffers A and B2 (B2: 20mM Tris, 500mM Imidazole pH7.4). After the collection of the middle peak fractions, the eluted material was filtered with 0.22 μm syringe filter and dialyzedagainst pre-chilled PBS (3.5L overnight at 4 °C and another 3.5L PBS for 4 hrs the next day) using 3.5 kDa MWCO dialysis membrane (Thermo Scientific 68035). Following another 0.22 μm syringe filtration, protein concentration was determined using a UV–VIS spectrophotometer with a theoretical extinction coefficient at 280 nm of 0.36 (mg/mL)^−1^ cm^−1^ . Protein was dispensed in single use aliquots and stored at −80 °C.

**Lab 2 Purification of recombinant WT α-synuclein**

Recombinant human WT α-synuclein protein was purified as previously reported [ref 59 in main manuscript]. Briefly, large culture flasks (1 L) of *E. coli* in LB (Millipore Sigma, Darmstadt, Germany; #L3022) containing antibiotics were grown at 37 °C while being shaken at 230 rpm using overnight auto induction kit ((Millipore Sigma, Darmstadt, Germany; #71300-4). The following day, the large culture was collected in 4 X 250ml beakers and pelleted. Each pellet were resuspended with 25 ml of lysis buffer (40% sucrose, 30 mM Tris HCl, 2 mM EDTA, pH 7.2 RT) and centrifuged at 9000× *g* at 20 °C for 20 min, the supernatant was discarded, and the pellet was resuspended with ice-cold Milli-Q H_2_O+2mM MgCl_2_.The suspension was centrifuged at 9000× *g* at 4 °C for 30 min. Supernatant was collected in a 150ml beaker to perform acid precipitation. The ph of the suspension was reduced to 3.5 using 1M HCL and the white precipitate was pelleted out by centrifuging at 9000× *g* at 4 °C for 30 min and supernatant collected, followed by increasing the ph back to 7.5 using 1M NaOH. Streptomycin sulfate 2.5 mg/mL was added to the suspension and left in rocker for 10 min at 4 °C, and were centrifuged for 20 min at 24,000× *g* at 4 °C. The supernatant was filtered using a 0.22-µm syringe filter and loaded through size exclusion chromatography column (Sephacryl S-200 HR 26/60 (Cytiva, Marlborough, MA, USA; #17119501) using NGC Chromatography System (Bio-Rad Laboratories, Inc., Hercules, CA, USA) and collected fractions from the target peak were then loaded into an anion exchange column (HiPrep Q FF 16/10 (Cytiva, Marlborough, MA, USA; #28936543). Chosen fractions were pooled and measured using at 280 nm using a NanoDrop spectrophotometer (Thermo Fisher Scientific, Waltham, MA, USA) using a 340-nm baseline correction, assuming an extinction coefficient of 5960 M^−1^ cm^−1^ and a molecular weight of 14.4 kDa. The monomer was diluted to yield a final concentration of <1 mg/mL using 40 mM sodium phosphate buffer pH 8 and stored at −80 °C.

The purified monomer was subjected to quality control methods such as SDS-PAGE, Western blot, Dynamic light scatter (DLS), MALDI-TOF Mass Spectrometry and CD Spectrum before using in SAA.

**Initial experiments by Lab 2 to tailor homogenization and assay conditions to the biopsies received, using samples unblinded to diagnosis**

**Salt titration of recombinant monomer**

A new batch of WT-αSyn recombinant monomer was purified, and a salt titration experiment was performed using SAA to detect the salt concentrations the monomer can withstand to perform efficiently without inducing spontaneous aggregation. Among the various concentrations tested, 40 mM NaCl was identified as optimal.

**Dilution factor**

Four samples (2 PD and 2 non-Lewy body disease negative controls) were used to optimize assay conditions. These samples were processed as mentioned in the methods section above and subjected to SAA using serial dilutions to detect an optimal and consistent SAA sample dilution factor that is detectable by the assay. A 1:10 dilution (10^-1^) was chosen as the optimal dilution factor after testing it on 5 assay runs.

**Assay duration**

Next, the assay duration was optimized by running the assay for a period of 84 h to account for spontaneous aggregation of the monomer, negative controls and the late aggregation of low-seeded positive controls. Combining these criteria, an assay duration of 42 h was used.

**Criteria for sample positivity**

The primary kinetic criterion used to label a sample positive was the number of replicates crossing the threshold. The threshold of 5,000 RFU was derived as µ+10σ, calculated from the first 10 readings of all samples (4 unblinded controls).

Samples were tested in quadruplicate and classified as 4/4, 3/4, 2/4, 1/4, or 0/4 well positive hits. When an SAA-tested sample scored a hit rate of 4/4 and 3/4, it was defined as positive without retesting. Samples scoring 2/4 were retested, whereby retested samples scoring ≥2/4 were defined as positive; otherwise, they were considered negative. Hit rates of 1/4 and 0/4 are retested with more dilutions to confirm that they remained ≤1/4 or 0/4 before labeling them as negative. This process helps make sure the low hit rates are not due to samples having high concentrations of seeds, which could inhibit seeding signals.

***Assessment for possible interference by keratin protein***

As cutaneous proteins such as keratin have beta-pleated sheet sequences, we tested our hypothesis that SAA of skin samples might non-specifically seed aggregation of synuclein monomers from non-Lewy body disease cases, decreasing apparent assay specificity. Skin biopsy positive controls from Lewy body disease cases were tested with and without spiked, commercially obtained keratin protein (Fig. 1), demonstrating no apparent keratin interference with specific

**Figure 1.** Synthetic keratin titration of skin biopsy homogenate SAA. A skin biopsy positive control homogenate (red) was tested with and without added keratin, titrated from 10mM to 500mM as quadruplicates. All replicates showed seeding signals with a uniform lag phase 0f 10 hrs demonstrating no apparent keratin interference with specific SAA seeding activity.

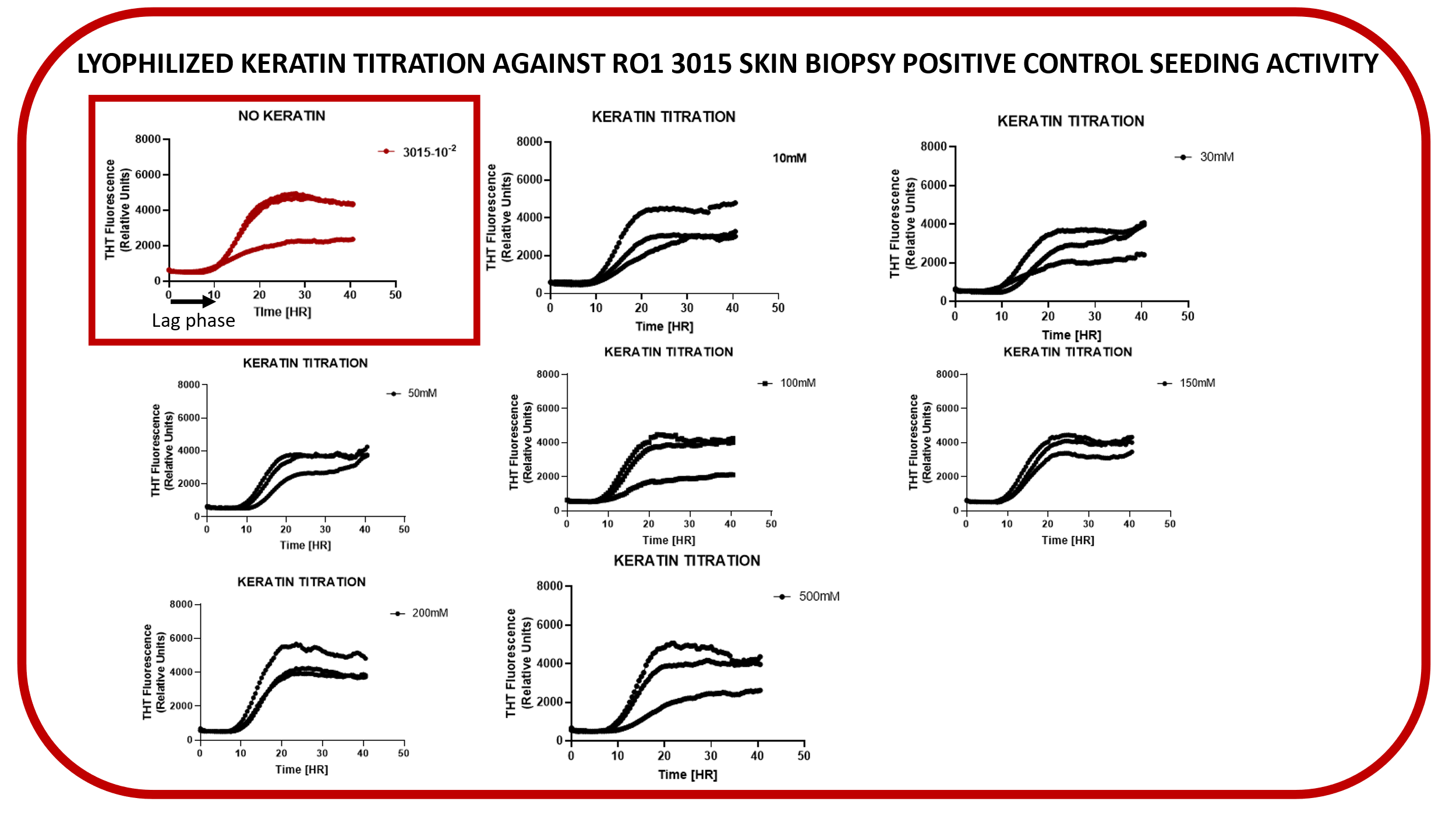
